# PSA and VIM DBS efficiency in essential tremor depends on distance to the dentatorubrothalamic tract

**DOI:** 10.1101/19013656

**Authors:** Till A. Dembek, Jan Niklas Petry-Schmelzer, Paul Reker, Jochen Wirths, Stefanie Hamacher, Julia Steffen, Haidar S. Dafsari, Mauritius Hövels, Gereon R. Fink, Veerle Visser-Vandewalle, Michael T. Barbe

**Author notes:** **Corresponding author:** Till A. Dembek, MD, Department of Neurology, University of Cologne, Kerpener Strasse 62, D-50837 Cologne, Germany. contributed equally (Till A. Dembek, Jan Niklas Petry-Schmelzer). **Funding:** This study did not receive additional funding.

## Abstract

**Objective:** To investigate the relation between deep brain stimulation (DBS) of the posterior-subthalamic-area (PSA) and the ventral-intermediate-nucleus (VIM) and the distance to the dentatorubrothalamic tract (DRTT) in essential tremor (ET).

**Methods:** Tremor rating scale (TRS) hemi-scores were analyzed in 13 ET patients, stimulated in both the VIM and the PSA in a randomized, crossover trial. Distances of PSA and VIM contacts to population-based DRTTs were calculated. The relationships between distance to DRTT and stimulation amplitude, as well as DBS efficiency (TRS improvement per amplitude) were investigated.

**Results:** PSA contacts were closer to the DRTT (p=0.019) and led to a greater improvement in TRS hemi-scores (p=0.005) than VIM contacts. Proximity to the DRTT was related to lower amplitudes (p<0.001) and higher DBS efficiency (p=0.017).

**Conclusions:** Differences in tremor outcome and stimulation parameters between contacts in the PSA and the VIM can be explained by their different distance to the DRTT.

## 1 Introduction

Essential tremor (ET) is the most common adult movement disorder, causes significant disability, interferes with activities of daily living, and reduces quality of life. For medication-refractory cases, deep brain stimulation (DBS) of the thalamic ventral intermediate nucleus (VIM) is an established, effective, and safe treatment.^1-3^ However, different groups have suggested that the posterior subthalamic area (PSA), which also incorporates the caudal zona incerta, might be an even more promising stereotactic target.^4-6^ In a recent prospective, randomized, double-blind crossover trial, our group compared stimulation in the VIM, the traditional DBS target, to stimulation in the PSA.^7^ PSA-DBS proved to be more efficient, yielding optimal stimulation effects at lower amplitudes with at least equivalent tremor control, while there was neither a quantitative nor qualitative difference in stimulation induced side effects between the two targets. However, the underlying neuroanatomical correlate remained unexplored.

DBS most likely modulates pathologic activity within the tremor network via cerebello-thalamo-cortical connections, i.e. the dentatorubrothalamic tract (DRTT).^8^ Direct targeting of the DRTT has been demonstrated to successfully control tremor^9, 10^ and effective contacts were located inside or close to the DRTT.^8^ However a direct correlation between clinical outcomes and the distance to the DRTT has not yet been proven.^8, 11^ We hypothesized that the observed differences between PSA and VIM stimulation depend on the proximity of contacts to the DRTT.

## 2 Methods

### 2.1 Study design

Ethics approval (Local vote 12-116), study registration (German Clinical Trials Register No. DRKS00004235), patient selection, implantation procedure and study design have already been described in detail.^7^ In brief, 13 patients with medication-refractory ET underwent bilateral stereotactic lead implantation. Landmark-based targeting was used to place leads with one contact on the intercommissural plane and at least one contact below in the PSA and one contact above in the VIM. Three months postoperatively patients entered a randomized, double-blind crossover to evaluate tremor improvement after two months of PSA-DBS and after two months of VIM-DBS. Current-controlled stimulation was used in all patients and stimulation pulse width was always set to 60 µs while frequencies ranged from 130 Hz to 200 Hz.

### 2.2 Clinical outcome and lead reconstruction

Tremor Rating Scale (TRS) hemi-scores (part A+B) were obtained at preoperative baseline, after two months of PSA-DBS, and after two months of VIM-DBS.^7^ For further analysis we calculated the DBS efficiency by dividing the TRS improvement from preoperative baseline by the stimulation amplitude (in mA). DBS leads were identified from postoperative CT scans and lead locations were transformed into the preoperative MRI using the Lead-DBS toolbox (www.lead-dbs.org).^12^

### 2.3 Average DRTT

Since the original study did not include individual diffusion imaging, we created one population-based averaged DRTT per hemisphere from high-quality diffusion imaging data of 32 healthy subjects of the Human Connectome Project (see acknowledgments), which consisted of multi-shell diffusion (bvals=1000, 3000, 5000; 256 directions) and T1 images. Brain extraction of the b0-images was performed using the BET-tool as implemented in the FMRIB software library (FSL) (FMRIB, Oxford, UK) and distributions of diffusion vectors were estimated for each voxel using BEDPOSTX.^13^ The number of fibers per voxel was set to three and diffusion coefficients were modelled using a Gamma distribution.^14^ Probabilistic fiber tracking was performed separately for each DRTT with PROBTRACKX2 using modified Euler integration.^13^ For all other parameters the respective default settings were used. The contralateral dentate nucleus was chosen as seed region, while the contralateral superior cerebellar peduncle, the ipsilateral red nucleus, and the ipsilateral precentral gyrus served as waypoints.^8, 9, 15, 16^ These regions of interest were defined in MNI space (ICBM 2009b NLIN asymmetric) by an experienced neurosurgeon (JW) and transformed to individual diffusion space using the SyN approach implemented in advanced normalization tools (ANTs, http://stnava.github.io/ANTs/;^17^ MNI to individual T1) and SPM 12 (http://www.fil.ion.ucl.ac.uk/spm/software/spm12/; individual T1 to diffusion space). The resulting fiber tracts were visually examined for anatomical accuracy, leading to 62 DRTTs included for further analysis. Each individual track frequency map was transformed into a track probability map according to Schlaier et al.^15^ and transformed back to MNI space. Finally, probability values per voxel were averaged over all subjects. The course of the population-based average DRTTs was validated in another cohort of nine essential tremor patients with individual diffusion imaging (see supplementary material).

#### Distance to average DRTT

For further analysis, the centers of gravity (COG) of each DRTT were calculated for each transversal slice as

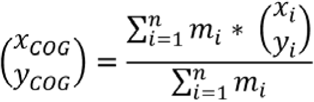

with *x*_*COG*_*/y*_*COG*_ representing the coordinates of the COG, and *m*_*i*_ representing the probability value of each voxel *i* with its coordinates *x*_*i*_ and *y*_*i*_ in the respective slice. The resulting coordinates were transformed from MNI space to the DBS patients’ individual T1 image via ANTs. Subsequently, the Euclidean distances between the right and left hemispheric VIM and PSA contacts and the closest COG of the respective average DRTT were calculated. The workflow is summarized in Figure 1.

**Figure 1.**
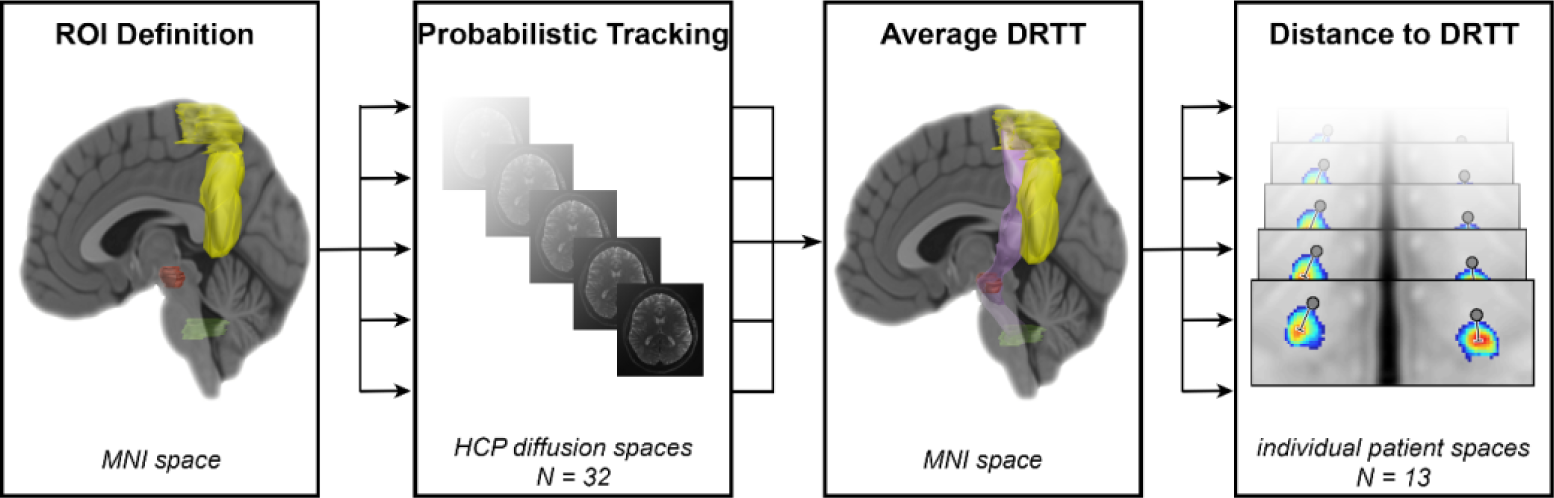
Workflow. **Legend:** Regions of interest (dentate nucleus=green, superior cerebellar peduncle, red nucleus=red, precentral gyrus=yellow) were defined in MNI space in both hemispheres and transformed to the diffusion space of each individual of the human connectome project dataset to perform probabilistic tracking. The resulting individual DRTTs were transformed back to MNI space and averaged to create one DRTT (lilac) per hemisphere. Finally the euclidean distance between each contact (grey dots) and the respective DRTT’s closest center of gravity (red hotspot) was calculated in the respective patient space. **Abbreviations:** DRTT=dentatorubrothalamic tract; HCP=Human Connectome Project; ROI=region of interest

### 2.4 Statistical analysis

TRS hemi-scores were analyzed with a linear mixed model for repeated measurements to account for the crossover design and multiple measurements per subject and per hemisphere.^7^ Differences in distance from the DRTT between PSA and VIM contacts were analyzed using the Wilcoxon signed-rank test. The relationship between distance to the DRTT and stimulation amplitude (in mA) as well as DBS efficiency were analyzed using linear mixed-effect models with the multiple measurements per subject as random-effect. Results are reported as (adjusted) mean values ± standard deviations. P-values p < 0.05 were considered statistically significant.

### 2.5 Data availability

Clinical data, stimulation parameters, lead locations, and averaged DRTTs are available via the Open Science Framework (OSF, dx.doi.org/10.17605/OSF.IO/MU827).

## 3 Results

Patient demographics, disease characteristics at baseline, and differences in stimulation parameters have been reported previously.^7^ Figure 2 illustrates the contact locations in relation to the DRTT. The adjusted means revealed better tremor suppression in the TRS hemi-score under PSA stimulation than under VIM stimulation (p=0.005, PSA: 4.54 (±1.19), VIM: 6.77 (±1.22); Figure 3A). The distance to the respective DRTT was shorter for PSA contacts (p=0.019, PSA: 2.13mm (±1.0), VIM: 2.81mm (±1.46); Figure 3B). Furthermore, the distance to the respective DRTT showed an effect on the stimulation amplitude (R^2^=0.73; p<0.001; Figure 3C) and on the efficiency of the stimulation site (R^2^=0.44; p=0.017; Figure 3D) with shorter distances indicating lower stimulation amplitudes and higher stimulation efficiency.

**Figure 2.**
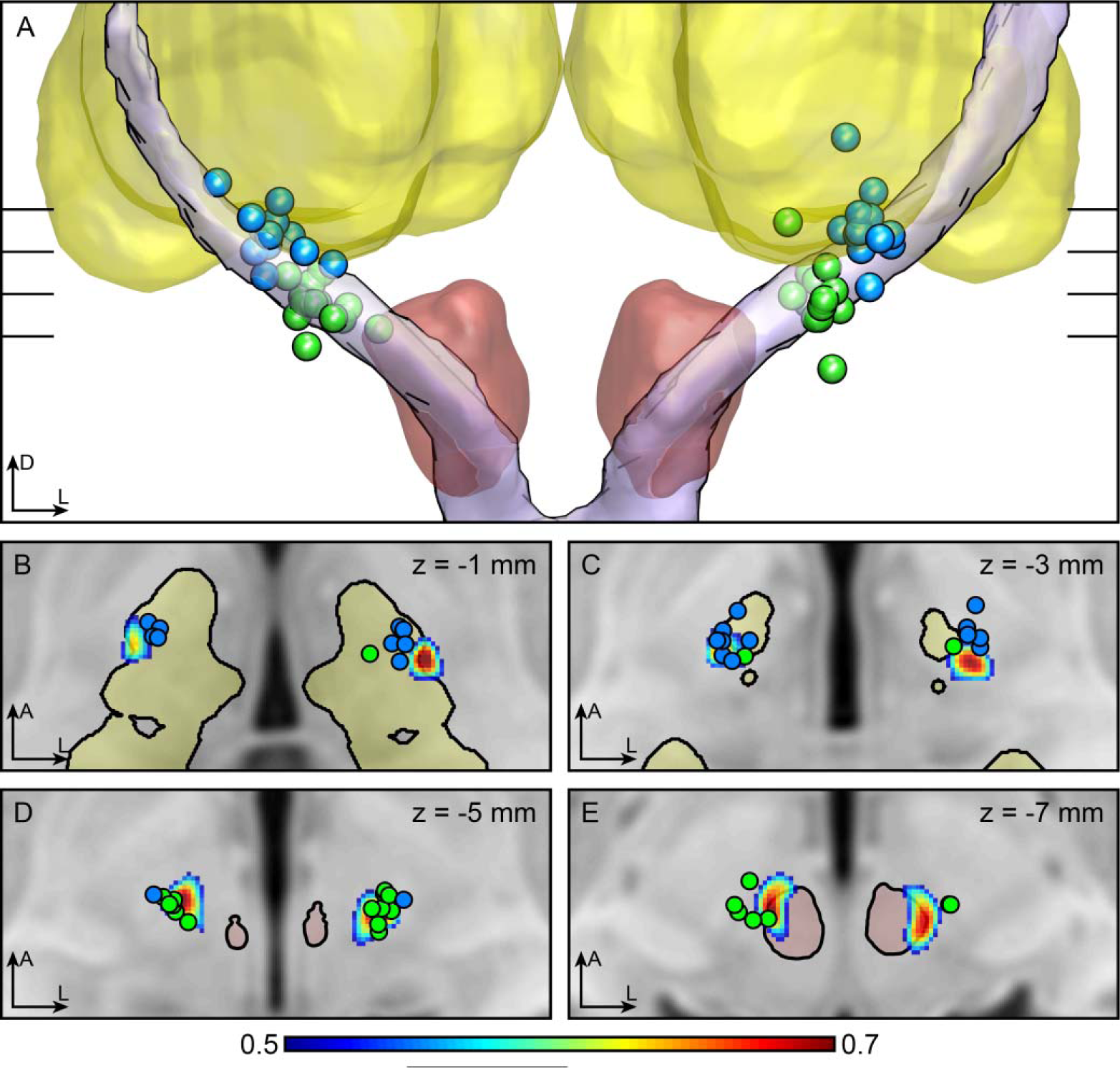
Contact positions. **Legend:** MNI-transformed contact positions (PSA=green, VIM=blue) in relation to the DRTT. A) 3D view from anterior with the red nucleus in red, the thalamus in yellow and the averaged DRTT in lilac.^18^ B-E) transversal slices through the T1 MNI template at the different heights indicated in (A). The DRTT is shown as probability map thresholded at 0.5 (see colorbar). **Abbreviations:** A=anterior; D=dorsal; L=left; DRTT=dentatorubrothalamic tract; PSA=posterior subthalamic area; VIM=ventral intermediate nucleus

**Figure 3.**
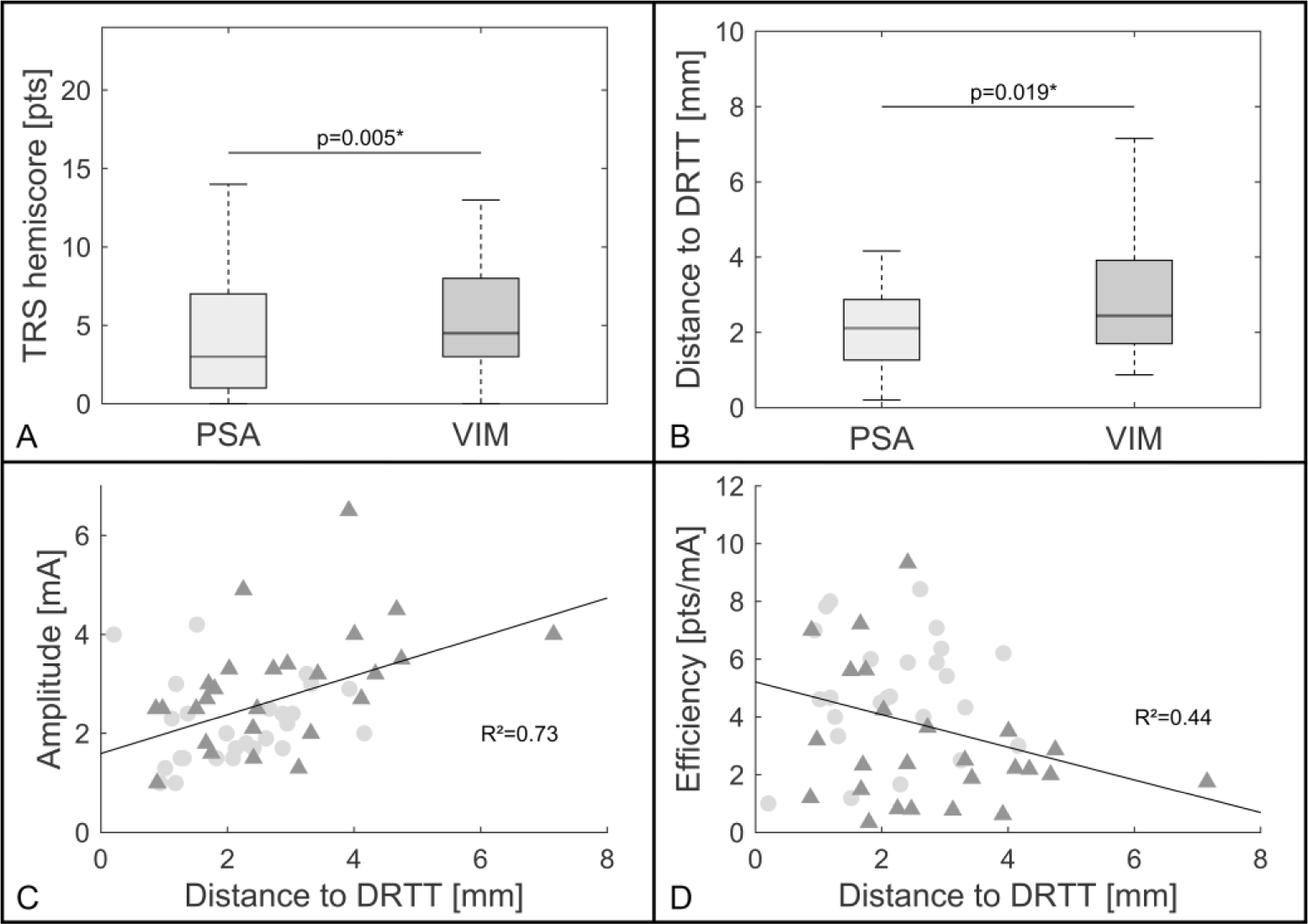
Results. **Legend:** Boxplots (median; interquartile interval; range) showing TRS hemi-scores (A) and the distance to the DRTT (B) for PSA and VIM stimulation. PSA stimulation led to lower TRS hemi-scores (p=0.005) and the distance to the DRTT was shorter for PSA contacts (p=0.023). C+D) Relationship between the distance to the DRTT of PSA (light circles) and VIM (dark triangles) contacts and the therapeutic stimulation amplitude (C) as well as the DBS efficiency (D). The shorter the distance to the DRTT, the lower the amplitude (p<0.001) and the higher the efficiency of the respective stimulation (p=0.018) as indicated by the dark line representing the fixed-effect. **Abbreviations:** DRTT=dentatorubrothalamic tract; PSA=posterior subthalamic area; VIM ventral intermediate nucleus

## 4 Discussion

This study demonstrates that the superior efficiency of PSA contacts in comparison to VIM contacts is related to their proximity to the DRTT. Distances to the center of the DRTT were significantly shorter in the PSA and this proximity lead to lower stimulation amplitudes and higher stimulation efficiency. So far, the evidence for a relation between stimulation of the DRTT and tremor improvement remained inconclusive.^8, 11^ One possible reason might be that the stimulation amplitude has not been considered as a crucial factor. While proximity to the DRTT seems to allow tremor control at low stimulation amplitudes, contacts farther from the DRTT could still achieve similar effects at higher amplitudes. Furthermore, several previous studies suggested that stimulation of the PSA or the caudal zona incerta might lead to at least equivalent^7, 19^ or even better tremor suppression than VIM-DBS.^5, 20^ In line with these studies, this post-hoc analysis indicates a superiority of the PSA by showing significant differences in TRS hemi-scores.

The major limitation of this study is that it did not include individual diffusion imaging. On the other hand, population-based^21^ or atlas-based^22^ DRTTs have been used to investigate DBS effects before. Additionally, the diffusion data used from the human connectome project is superior in quality to most images acquired during clinical routine and while a population-based average tract might neglect individual neuroanatomic aberrations, it might also be less prone to tracking errors which have been shown to occur when identifying individual DRTTs.^15^ Most importantly, the average deviation of individual patient DRTTs from our population-based DRTT was within the range of image accuracy (see supplementary material). We employed probabilistic fiber tracking, as it might be better in detecting the DRTT than the deterministic algorithms embedded in commercially-available stereotactic planning software.^15^

Even though landmark-based targeting was used when implanting our patients (PSA vs. VIM)^7^, the results nonetheless could be explained by proximity to the DRTT. In conclusion, direct targeting of the DRTT, which has been demonstrated to be feasible, could improve outcome even further.^9^ Future studies, which prospectively compare direct targeting of the DRTT to landmark-based targeting are needed. Moreover, it should be explored whether visualization of the DRTT can also guide postoperative DBS programming. With population-based averaged tracts this could even be investigated in patients who have not received individual diffusion imaging prior to implantation.

## Abbreviations

DBS: deep brain stimulation
DRTT: dentatorubrothalamic tract
ET: ssential tremor
PSA: posterior subthalamic area
VIM: ventral intermediate nucleus

## Acknowledgement

Data collection and sharing for this project were provided by the Human Connectome Project (HCP; principal investigators: Bruce Rosen, MD, PhD, Arthur W. Toga, PhD, Van J. Weeden, MD). HCP funding was provided by the NIH National Institute of Dental and Craniofacial Research, National Institute of Mental Health, and National Institute of Neurological Disorders and Stroke. HCP data are disseminated by the Laboratory of Neuro Imaging at the University of Southern California. HCP is the result of efforts of coinvestigators from the University of Southern California, Martinos Center for Biomedical Imaging at Massachusetts General Hospital, Washington University, and University of Minnesota.

## Author disclosures for the preceding 12 months

Till A. Dembek reports speaker honoraria from Boston Scientific and Medtronic.

Jan Niklas Petry Schmelzer reports a travel grant from Boston Scientific.

Paul Reker reports a travel grant from AbbVie.

Jochen Wirths reports a travel grant by Boston Scientific.

Stefanie Hamacher has nothing to disclose.

Julia Steffen reports a travel grant from BIAL.

Haidar S. Dafsari reports funding from the Prof. Klaus Thiemann Foundation, the Koeln Fortune Program, and the Felgenhauer Foundation and has received speaker honoraria from Boston Scientific and Medtronic

Gereon R. Fink has nothing to disclose.

Mauritius Hoevels has nothing to disclose.

Veerle Visser-Vandewalle is a member of the advisory boards and reports consultancies for Medtronic, Boston Scientific and Abbott and received a grant from SAPIENS Steering Brain Stimulation (now Medtronic Eindhoven Design Center).

Michael T. Barbe reports speaker honoraria from Medtronic, Abbott, GE Medical, UCB, Bial and research grants from Medtronic.

## Author contributions

1. Research project: A. Conception, B. Organization, C. Execution; 2. Statistical Analysis: A. Design, B. Execution, C. Review and Critique; 3. Manuscript: A. Writing of the first draft, B. Review and Critique.

T.A.D.: 1A-C, 2A-C, 3A-B

J.N.P.S.: 1A-C, 2A-C, 3A-B

P.R.: 1C, 2C, 3B

J.W.: 1C, 2C, 3B

S.H.: 2A-C, 3B

J.S.: 1C, 2C, 3B

H.S.D.: 1C, 2C, 3B

M.H.: 1C, 2C, 3B

G.R.F.: 2C, 3B

V.V.V.: 1C, 2C, 3B

M.T.B.: 1A, 1C, 2C, 3b

## Notes

**Financial disclosures/ conflict of interest concerning the research related to the manuscript:** None.

### Competing Interest Statement

The authors have declared no competing interest.

